# Using machine learning to improve the readability of hospital discharge instructions for heart failure

**DOI:** 10.1101/2023.06.18.23291568

**Authors:** Alyssa W. Tuan, Nathan Cannon, David Foley, Neha Gupta, Christian Park, Kyra Chester-Paul, Joanna Bhasker, Cara Pearson, Avisha Amarnani, Zachary High, Jennifer Kraschnewski, Ravi Shah

## Abstract

**Background:** Low health literacy is associated with poor health outcomes. Hospital discharge instructions are often written at advanced reading levels, limiting patients’ with low health literacy ability to follow medication instructions or complete other necessary care. Previous research demonstrates that improving the readability of discharge instructions reduces hospital readmissions and decreases healthcare costs. We aimed to use artificial intelligence (AI) to improve the readability of discharge instructions.

**Methodology/Principal Findings:** We collected a series of discharge instructions for adults hospitalized for heart failure (n=423), which were then manually simplified to a lower reading level to create two parallel sets of discharge instructions. Only 343 sets were then processed via AI-based machine learning to create a trained algorithm. We then tested the algorithm on the remaining 80 discharge instructions. Output was evaluated quantitatively using Simple Measure of Gobbledygook (SMOG) and Flesch-Kincaid readability scores and cross-entropy analysis and qualitatively. Using this test dataset (n=80), the average reading levels were: original discharge instructions (SMOG: 10.5669±1.2634, Flesch-Kincaid: 8.6038±1.5509), human-simplified instructions (SMOG: 9.4406±1.0791, Flesch-Kincaid: 7.2221±1.3794), and AI-simplified instructions (SMOG: 9.3045±0.9531, Flesch-Kincaid: 7.0464±1.1308). AI-simplified instructions were significantly different from original instructions (p<0.00001). The algorithm made appropriate changes in 26.1% of instances to the original discharge instructions and improved average reading levels by 1.26±0.32 grade levels (SMOG) and 1.02±0.47 grade levels (Flesch-Kincaid). Cross-entropy analysis showed that as the data set increased in size, the function of the algorithm improved.

**Conclusions/Significance:** The AI-based algorithm learned meaningful phrase-level simplifications from the human-simplified discharge instructions. The AI simplifications, while not in complete agreement with the human simplifications, do appear as statistically significant improvements to SMOG and Flesch-Kincaid reading levels. The algorithm will likely produce more meaningful and concise simplifications among discharge instructions as it is trained on more data. This study demonstrates an important opportunity for AI integration into healthcare delivery to address health disparities related to limited health literacy and potentially improve patient health.

**Author summary:** Patient-facing materials are often written at too high of a reading level for patients, such as hospital discharge instructions. These instructions provide critical information on how to control health conditions, take medications, and attend follow-up visits. Difficulty understanding these instructions could lead to the patient returning to the hospital if they do not understand how to control their health condition.

Improving the readability of discharge instructions can reduce hospital readmissions. It may improve health outcomes for patients and reduce healthcare costs. Artificial intelligence (AI) may be used to improve the reading level of patient-facing materials. Our work aims to create a tool that can accomplish this goal.

We obtained hospital discharge instructions for heart failure. Discharge instructions were edited by medical experts to improve their readability. This created two sets of discharge instructions that were processed using AI. We created and tested an AI tool to automatically simplify discharge instructions. Although not perfect, we found that the tool was successful. This research shows that AI can be used to address health literacy needs within health care by making patient-facing health materials easier to understand. This is important to empower all patients to take action to improve their health.

## Introduction

Limited health literacy is an important predictor of health status and affects about 9 out of 10 adults in the United States **[1]**. It is defined as the degree to which individuals can find, understand, and use information and services to inform health-related decisions and actions for themselves and others **[2]**. Limited health literacy is related to health disparities **[3-7]**, higher healthcare costs **[8-14]**, and poorer health outcomes **[15]**.

Patient-facing literature is often written at too high of a readability level for their intended recipients **[16]**, including hospital discharge instructions **[13-14, 17]**. This can make it difficult for patients to understand their medication dosing instructions, timeline for follow-up appointments, and other aspects of care. As a result, patients could be readmitted to hospitals **[18, 19]**, resulting in increased costs for the healthcare system as a whole with the risk of increasing morbidity and mortality for patients. A study by Choudhry et al. demonstrated improved readability of discharge summaries by breaking up long sentences, changing complex terminology, and assessing content with readability calculators. As a result, the study team found that 30-day patient readmissions in the post-hospital setting were reduced by 50% **[20]**.

Our study aims to leverage natural language processing and artificial intelligence (AI) to improve the readability of hospital discharge instructions. AI can be used to address contextual information, grammatical structure, and changes to word order. Most current AI solutions related to the discharge process revolve around care coordination **[21]**, rather than focus on patient discharge instructions, or aim to predict patients likely to be readmitted to the hospital **[22-24]**. We examined whether an AI-based system can learn and incorporate meaningful simplifications of hospital discharge instructions. We believe that we were able to achieve this aim.

## Materials and methods

### Study overview

Institutional Review Board approval was obtained for this study. We obtained a list of medical record numbers for adults hospitalized from 2016 to 2021 at our academic medical center with a diagnosis code of heart failure, a leading cause of readmissions, addressed during their hospitalization. This list was obtained through our institution’s information management department. Discharge summaries for these hospitalizations written between 2016 to 2021 were then collected manually from the electronic medical records.

We collected 423 discharge instructions. These discharge instructions were not randomly collected from the list of medical record numbers available. Collected discharge instructions were re-formatted to replace bulleted lists with sentences. This ensured that bulleted lists were not perceived as run-on sentences by readability calculators. The readability scores of the discharge instructions were then calculated using the Simple Measure of Gobbledygook (SMOG) index and Flesch-Kincaid score **[25]**, creating a compilation of reading level scores for the original set of discharge instructions that correlate to years of education and grade reading level.

The original versions of discharge instructions were then manually rewritten by medical experts as human simplifiers using a standardized process to improve readability. This process included eliminating long sentences and exchanging complex terms with lay terms. The manual simplification of text was performed on a line-by-line basis to create parallel data sets for AI training, while working to preserve the original meaning of the content for patients. The readability scores of the human-simplified discharge instructions were then calculated using the SMOG index and Flesch-Kincaid score.

Of the 423 discharge instructions, only 343 parallel discharge instructions (“training dataset”) were processed via AI-based machine learning to create an algorithm trained on the data. This algorithm was then tested on the remaining 80 discharge instructions that the algorithm had not seen before (“test dataset”). The output from the algorithm was evaluated quantitatively using readability scores and cross-entropy analysis, which measures how well the algorithm predicts human simplification on a word-by-word basis. The output was also evaluated qualitatively by human readers examining grammar and semantic content.

### Algorithm development

The text-simplification task is formulated as a machine translation problem. Machine translation algorithms take text in a source language and output text in a target language, such as English or German. In our case, the source language is text from the original versions of hospital discharge instructions, and the target language is an AI-simplified version of the text. "Translation" in this case does not refer to translation between different languages, but rather the formal operation of the algorithm. All text, both input and output, is rendered in English.

Like most state-of-the-art machine translation algorithms, our model is built on transformers. Transformers are neural network architectures that learn representations of input sequences from attention-based transformations.

## Results

The average SMOG reading level of the training dataset (n=343) with no simplifications was 10.9300 ± 1.4272. By Flesch-Kincaid, the average reading level was 9.0776 ± 1.7771. The average reading level of the human-simplified instructions was 9.3110 ± 0.8953 by SMOG and 7.3081 ± 1.2092 by Flesch-Kincaid.

We found that the average SMOG reading level of the test dataset (n=80) was 10.5669 ± 1.2634 for the original discharge instructions and 9.4406 ± 1.0791 for human-simplified instructions (**Fig 1**). The average Flesch-Kincaid reading level was 8.6038 ± 1.5509 for the original discharge instructions and 7.2221 ± 1.3794 for human-simplified instructions,

**Fig 1:**
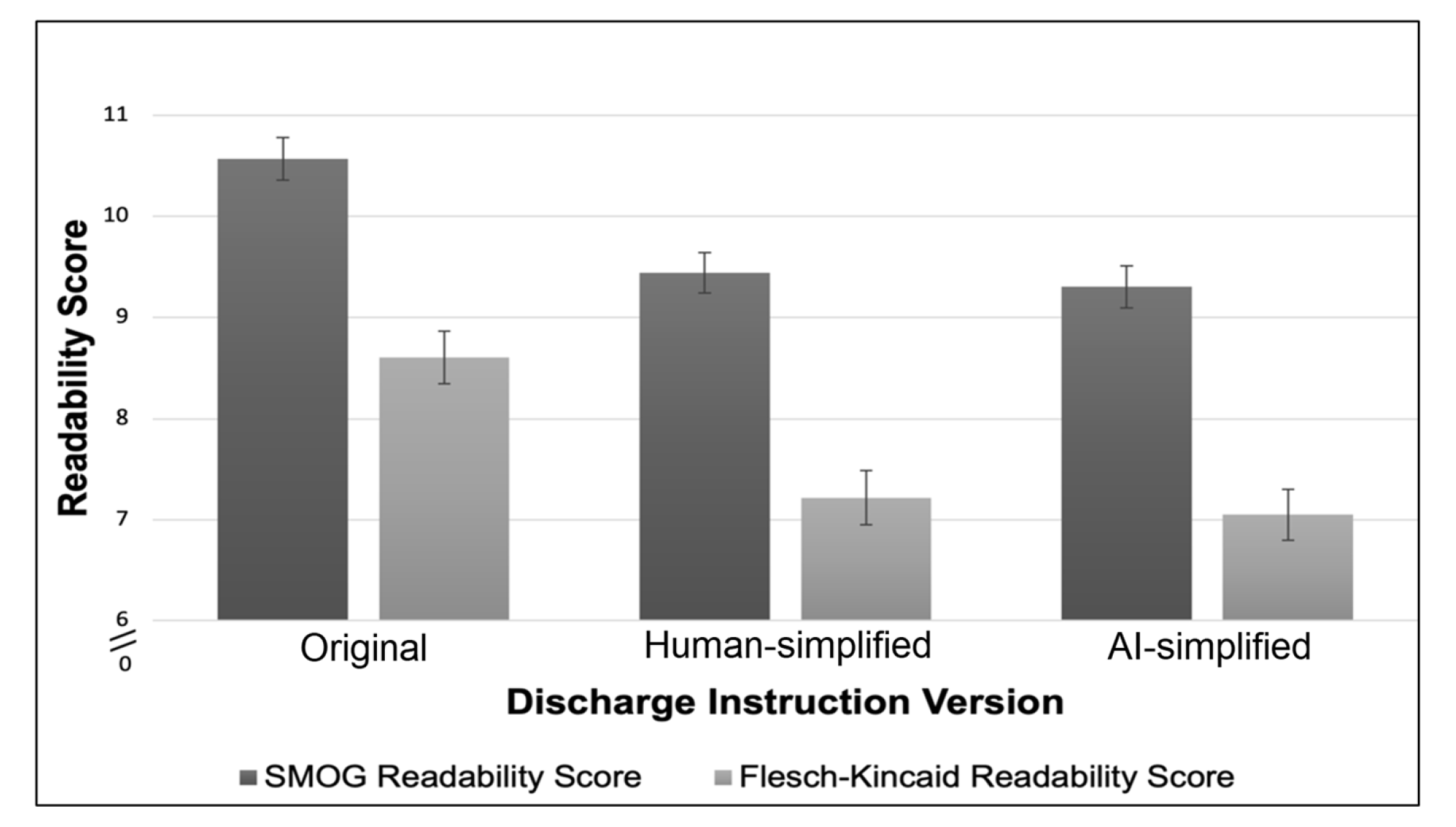
Average readability scores for the three versions of discharge instructions. Average readability scores of the original, human-simplified, and AI-simplified discharge instructions (n=80) using the SMOG index and Flesch-Kincaid readability scores.

In comparison, the average SMOG reading level for the AI-simplified instructions in the test dataset (n=80) was 9.3045 ± 0.9531 (**Fig 1**). The human-simplified and the AI-simplified were both significantly improved reading levels from the original discharge instructions (**Table 1**). The average Flesch-Kincaid reading level was 7.0464 ± 1.1308 for AI-simplified instructions. We found that the algorithm made appropriate changes to the original discharge instructions in 26.1% of instances (**Table 2**), as well as average readability level improvements of 1.2624 ± 1.0791 grade levels by the SMOG index and 1.5574 ± 1.3274 grade levels by the Flesch-Kincaid score. We did find that the algorithm may maintain the original text without making any simplifications, although a human simplifier could identify opportunities to simplify the text. At times, the algorithm would introduce text that did not make sense within the meaning of the sentence (**Table 3**). The cross-entropy analysis showed that mean cross-entropy decreased as the data set increased in size, indicating that the function of the algorithm improved as the data set size grew (**Fig 2**).

**Fig 2:**
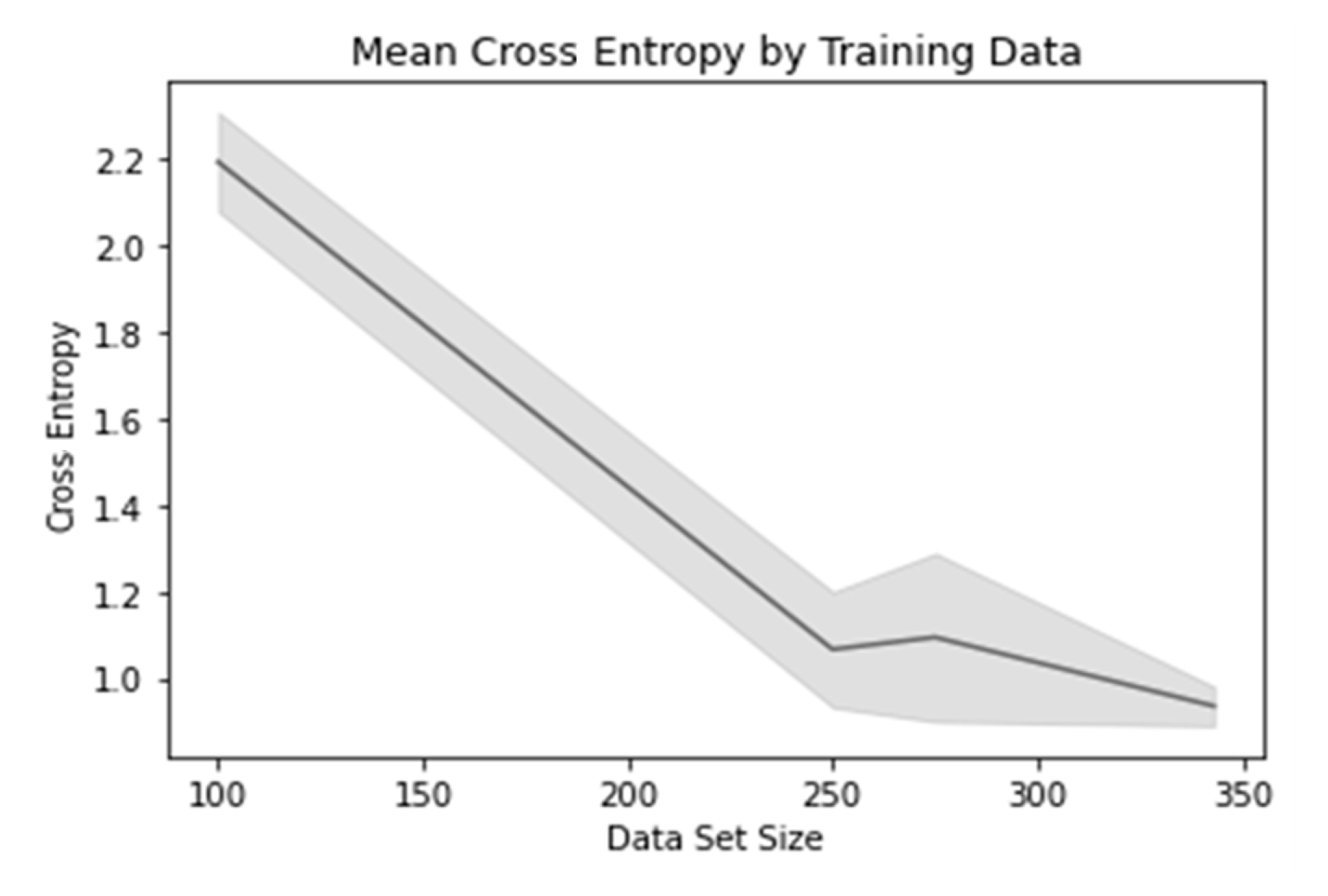
Mean cross entropy by the size of the hospital discharge instruction data set. Cross entropy decreased as the data set increased in size, indicating that the algorithm improved as the size of the data set grew (n=343).

**Table 1:**
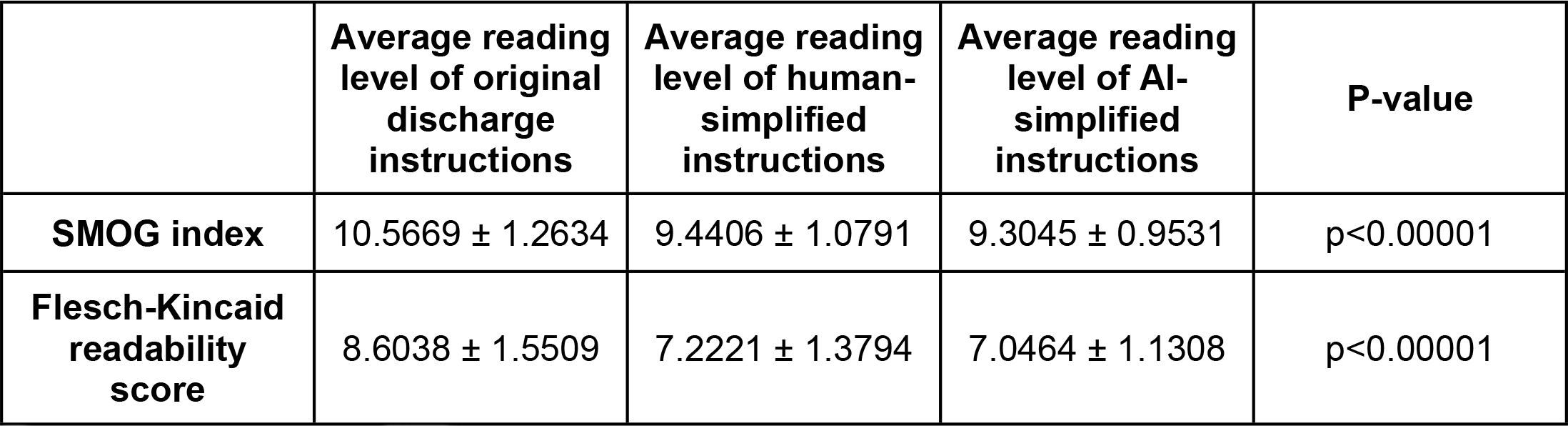
Comparing average readability scores among the three versions of discharge instructions (n=80).

**Table 2:** Examples of text from original discharge instructions with correlating human-simplified and AI-simplified text.

| Original discharge instructions | Human-simplified text | AI-simplified text |
| --- | --- | --- |
| Please take all of your medications exactly as prescribed. Tell your primary care provider if you cannot afford your medications. | Please take all of your medicine exactly as <u>written</u>. Tell your <u>family doctor</u> if you <u>can't pay for</u> your <u>medicine</u>. | Please take all of your medications exactly as <u>ordered by the doctor</u>. Tell your <u>family doctor</u> if you can not <u>pay for</u> your medications . |
| It was our pleasure to care for you during your hospitalization. | It was our pleasure to care for you during your <u>stay in the hospital</u>. | It was our pleasure to care for you during your <u>hospital stay</u>. |
| If you find a discrepancy between your home medications and the medication list found later in this packet, please address this with your family doctor. | If you find a <u>difference</u> between your <u>home drugs</u> and the <u>drug</u> list found later in this packet, please <u>ask about</u> this with your family doctor. | If you find a <u>difference</u> between your home medications and the medication list found later in this packet , please <u>talk about</u> this with your family doctor . |

**Table 3:** Examples of text from original discharge instructions with correlating human-simplified and AI-simplified text, with AI-simplified text unchanged or inappropriately simplified.

| Original discharge instructions | Human-simplified text | AI-simplified text |
| --- | --- | --- |
| This number will connect you to the hospital operator and ask for the Cardiac Surgery PA on duty. | This number will get you to the hospital operator and ask for the <u>heart surgeon</u> on duty. | This number will connect you to the hospital operator and ask for the <u>Cardiac Surgery PA</u> on duty. |
| If you do not hear from the provider on this date, please contact them on the next business day. | If you do not hear from the <u>doctor</u> on <u>that day</u>, please call them the next <u>weekday</u>. | If you do not hear from the <u>doctor</u> on this date , please contact them on the next <u>2</u> business day. |
| Do not discontinue medications without first discussing with your physician. | Do not <u>stop</u> medications without first discussing with your <u>doctor</u>. | Do not <u>put any first pee first</u> discussing with your doctor. |

## Discussion

We found that our AI-based algorithm learned meaningful phrase-level simplifications from the human-simplified discharge instructions. The AI simplifications, while not completely agreeing with the human simplifications, do appear as statistically significant improvements to SMOG and Flesch-Kincaid reading levels. Future studies should evaluate patient comprehension of the simplified texts. A similar study identified difficult terms, replaced them with easier synonyms, and generated and inserted explanatory texts for them **[26]**. Most text replacements were deemed correct and user evaluation showed a non-statistically significant trend toward better comprehension when translation was provided. Another study automated simplification of medical text based on word frequencies and language modeling using medical ontologies enriched with lay terms **[27]**. The language model was trained on medical forum data and tested using crowdsourcing. The researchers found their model generated simpler sentences while preserving grammar and the original meaning. One study, however, used natural language processing to substitute difficult terms for simpler terms and split long sentences into shorter sentences in electronic medical records and journal articles; it found that the length of sentences and reading grade level increased from baseline; however, the study did not evaluate comprehension **[28]**.

Our study’s cross-entropy findings show that the algorithm is more concise and does not produce unnecessarily long sentences. Decreasing cross entropy with increased data set size indicates that the AI-based algorithm will likely produce more meaningful and concise simplifications among discharge instructions as we continue to train the algorithm on more data sets. Studies investigated methods to expand resources that link medical terms with lay terms, which may facilitate algorithm training. These methods include creating a system that identifies terms based on their importance for patients **[29]**, ranking medical terms mined from electronic medical records by importance for patient comprehension **[30]**, and predicting which medical terms are unlikely to be understood by a lay reader **[31,32]**.

Text simplification via machine learning can be a challenging process, but it serves an important role in reducing barriers to health literacy. Health literacy is associated with a range of social and individual factors **[3-6]**, and certain populations are likely more adversely affected by limited health literacy compared to others without such limits, such as certain racial groups **[4]**, seniors **[6]**, young adults **[6]**, and Medicare and Medicaid populations **[7]**. Limited health literacy is associated with increased risk of hospitalization **[33,34]**, mortality **[35]**, and high healthcare costs **[8,9]**. High healthcare costs could be driven in part by individuals’ difficulty understanding how to manage their chronic conditions, hospital discharge instructions **[10-14]**, and/or other medical-related literature.

There are opportunities to assess patients’ health literacy levels to determine their literary needs. A study found that only 20% of hospitals reported routinely screening patients and 41% of hospitals reported never screening health literacy **[36]**. Hospitals that screen for health literacy may have higher medication adherence **[36]** and reduced hospital readmission rates **[37]**. Screening tools to measure health literacy include the Brief Health Literacy Screen (BHLS) **[38]**, Test of Functional Health Literacy (S-TOFHLA) **[38,39]**, Single Item Literacy Screener (SILS), and Newest Vital Sign (NVS). Other opportunities to address health literacy include tailoring education **[40]**, engaging caregivers **[41]** and conducting activities that increase patients’ self-efficacy **[42]**.

This study is limited by its small sample size of hospital discharge instructions, as there is a lack of high-quality, medically accurate, and publicly available datasets for evaluating medical text simplification. These discharge instructions were not randomly collected from the list of medical record numbers available. Further, our study is limited to heart failure discharge instructions, which may have less variability than those for other diagnoses. Expanding our data set is a priority for future studies, as well as evaluating comprehension of AI-simplified discharge instructions.

AI has the potential to improve the lives of individuals. This includes within improving the readability of health-related materials. There is a need to ensure an appropriate match between the readability of the health content and the health literacy level of the patient. The association of limited health literacy with poor outcomes such as increased risk of hospitalization and death warrants the need for continued investigations into such interventions.

## Data Availability

The code used for algorithm training is publicly available at https://www.kaggle.com/code/daveyfoley/readability-transformer. The SNOMED ontology used in the development of the algorithm can be accessed at snomedct.org. Because SNOMED is protected by copyrights, elements of the code involving SNOMED, as well as the algorithm itself, cannot be shared publicly. Out of concern for the protection of patient privacy and confidentiality, we are unable to make the full training dataset publicly available. However, a deidentified partial training set and data analysis can be found at kaggle.com/datasets/ntcannon/SimplifyTrainingDataset. Additionally, access to the full dataset and algorithm is available to researchers who meet the criteria for access to confidential data and can be obtained by contacting the Penn State Institutional Review Board or corresponding author, Nathan Cannon, at.

## Acknowledgements

We thank the Institute for Augmented Intelligence in Medicine (I.AIM) at Northwestern University that hosted the First Annual Big Ten Augmented Intelligence Bowl, which served as the initial stimulus for this research project.

